# Textiloma trends in Colombia: Ecological analysis 2015 – 2024

**DOI:** 10.1101/2025.11.06.25339712

**Authors:** Sharit Lineht Quevedo Rodríguez, Janis Rivera Díaz, Ximena Daniela Zabaleta Bello, Wanderley Augusto Arias-Ortiz

## Abstract

The analysis of textiloma behavior during the study period revealed marked territorial and sex-related heterogeneity. Departments such as Putumayo, Magdalena, Meta, and Quindio showed the highest averages among men, whereas regions like the Archipelago of San Andres and Guaviare presented outlier values among women. These differences reflect an unequal distribution of risk and variability in surgical and control practices, underscoring the need to strengthen prevention mechanisms. Preventing textilomas is a key indicator of patient safety and the quality of surgical practice. It requires concrete actions such as strict adherence to safe surgery protocols, systematic verification of counts, traceability of surgical materials, and continuous training of healthcare personnel in a culture of safety. Furthermore, the incorporation of intelligent analysis and traceability systems could enhance institutional surveillance, allowing the transformation of collected data into useful evidence for decision-making and the design of public policies aimed at reducing the incidence of this adverse event.

## Introduction

A *textiloma* is a mass formed by textile material (compresses, towels, or gauze) accidentally retained in the body after surgery, capable of causing a foreign body reaction and forming a granuloma that can be mistaken for a tumor or abscess (1). According to the Royal Spanish Academy (RAE), this term, also known as “oblito,” is used to describe the forgetting of a foreign object in the patient’s body after performing a surgical procedure (2), considered an adverse (sentinel) and preventable event, as it can lead to complications such as infections, chronic pain, obstructions, and even put the patient’s life at risk associated with medical negligence (3).

Globally, studies estimate that the incidence of textiloma is between 1 in 1,000 and 1 in 10,000 surgical procedures performed (4). According to the Ministry of Health and Social Protection (MSPS) in the 2009 technical guide “Good practices for patient safety in health care” for Colombia, the frequency of textile foreign bodies is higher than the officially reported figures, as there are more cases of forgotten foreign objects than are actually reported (3). In Colombia, information on textilomas comes mainly from internal hospital reports and adverse event records from the Mandatory Health Quality Assurance System (SOGCS), without a consolidated national epidemiological characterization (4).

The Technical Guide to Good Practices for Patient Safety and Safe Care, issued by the Ministry of Health, establishes guidelines and processes aimed at preventing adverse events and ensuring quality care (5). It includes evidence-based recommendations, instructional packages for healthcare personnel, and mandatory standards ranging from proper patient identification and effective communication to infection prevention, safe use of medications, and safety in surgical procedures. It also promotes an institutional culture of safety and patient participation in their own care (5).

Preventing medical errors is a priority in patient safety, and to avoid them, in the surgical context, there are rigorous protocols, including the WHO surgical checklist, counting all instruments placed on the operating table and counting all white material before starting the procedure, during the procedure, and before closing the surgical field, as well as effective information sharing among the surgical team (3).

Understanding the prevalence and behavior of textilomas in patients after surgery is essential to gauge the true magnitude of this adverse event (6). Identifying risk factors and recognizing patterns of occurrence (such as type of surgery, type of emergency, time of procedure, and duration) allows for the evaluation of the effectiveness of safety protocols and the reinforcement of surgical care systems in hospital institutions (5,7).

In this context, surgical items left behind in patients are not considered completely avoidable, but they are preventable (8). Despite professional opinions and case reports, few studies offer quantitative information at the national level regarding the discovery of foreign bodies accidentally left behind in body cavities.

However, studies have been documented in other countries, such as Uruguay, where a study of seven patients who had forgotten sponges after surgery was found. Two were asymptomatic and five symptomatic (two early, three late) (7). In Venezuela, the study mentions the most frequent elements found in cavities, which are: sponges 69%, instruments 31%, clamps 7%, and others 24% (9). Both articles refer to failures in the attention process and reinforce the approach of public policies related to patient safety and the lack of communication within the surgical team.

The present study sought to describe the distribution of the number of people served for textilomas accidentally left in body cavities in Colombia for the period 2015-2025, identifying statistical differences between departments and years of occurrence. In addition, it analyzed the distribution of textiloma throughout the study period.

## Methodology

An observational and retrospective study was conducted, based on secondary sources, by consulting the Individual Health Services Provision Registry (RIPS) for the number of people treated with diagnosis T81.5 “foreign body accidentally left in body cavity or surgical wound following procedure” in institutional care reports in Colombia.

The search was conducted on August 2^nd^, 2025, and included records with the code T81.5 of the International Classification of Diseases (ICD-10), corresponding to the period 2014-2024. All case reports designated as new confirmed and repeated cases of textiloma were included.

Information was collected considering the sociodemographic characteristics of patients, the attention department, cases reported by department, health system, and sex, to identify possible patterns and variations in the occurrence of these cases according to the geographic and population context.

### Analysis plan

To analyze the behavior between new confirmed cases and repeat confirmed cases, a case analysis was performed comparing their behavior over the years, for which a graphical method was used. Prevalence rates per million inhabitants were calculated. Rates were compared by the attention department, health insurance system and sex.

The analysis of behavior over time was performed using a graphical method to analyze trends and variations, which included calculating the year-on-year percentage variation and for the period. To establish the trend, a simple linear regression was applied, and the model performance was analyzed by the slope and R2 coefficient. In addition, measures of central tendency and dispersion were applied to understand the variability between departments each year and between years by geographic unit. This made it possible to determine whether there was homogeneity in the behavior of both analyses.

On the other hand, choropleth maps were designed to illustrate the geographical distribution of prevalence across the departments. The Z-test for independent rate comparison was applied to analyze differences between sex by year. To evaluate the independence between sex and health insurance system, Pearson’s chi-square test was applied. The level of statistical significance was 0.05. MS Excel 365 was used for statistical analysis, hypothesis testing was performed with Epidat 3.1, and maps were created with ArcGis Pro. In addition, the incidence ranking and chart by department and the box plots were created using the fluorish tool. Generative Artificial Intelligence was used in some parts of the manuscript to enhance the clarity and precision of the English language from the original draft, always under full human oversight and intellectual control. All conceptual and analytical content was developed entirely by the authors.

## Results

A total of 2,092 cases were identified during the period from 2015 to 2024, of which 69.12% (N=1,446) were new cases and 30.87% (N=646) were repeat cases.

In 2015, the total number of cases remained relatively stable, with 97 cases reported, increasing to 118 in 2016. On the other side, a steady increase was observed in 2018 and 2019, with a peak of 141 cases.

Referring to the new cases, it is observed that they were predominant throughout the entire period analyzed, with values ranging from 44 to 95 on average. In addition, there was continuous growth during the study period.

Repeat cases showed a different pattern. Between 2015 and 2016, reports remained at a lower level compared to new cases, with values between 26 and 30 cases. They then increased significantly between 2017 and 2019, reaching a peak of 49–50 cases (Figure 1)

**Graphic 1.**
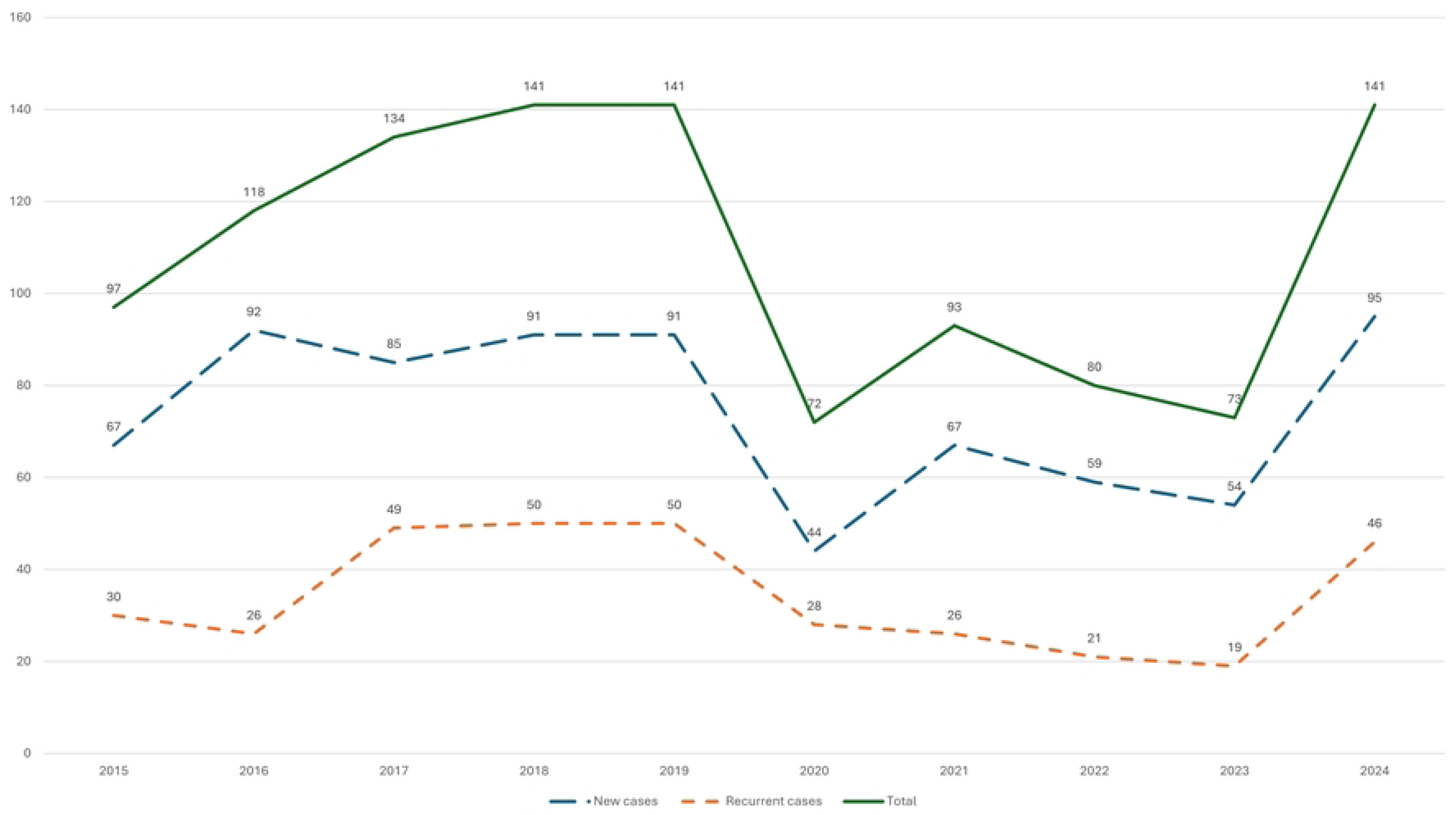
Trend Analysis of textiloma: New and repeated cases for the period (2015–2024).

In terms of the analysis by sex between 2015 and 2024, the prevalence rates of textiloma in Colombia varied throughout the period. In the early years, rates remained between 1.2 and 2.0 per million inhabitants, with similar values between males and females (2017– 2019; p > 0.8). In 2020, there was a decrease in both men and women.(Figure 2).

**Figure 2.**
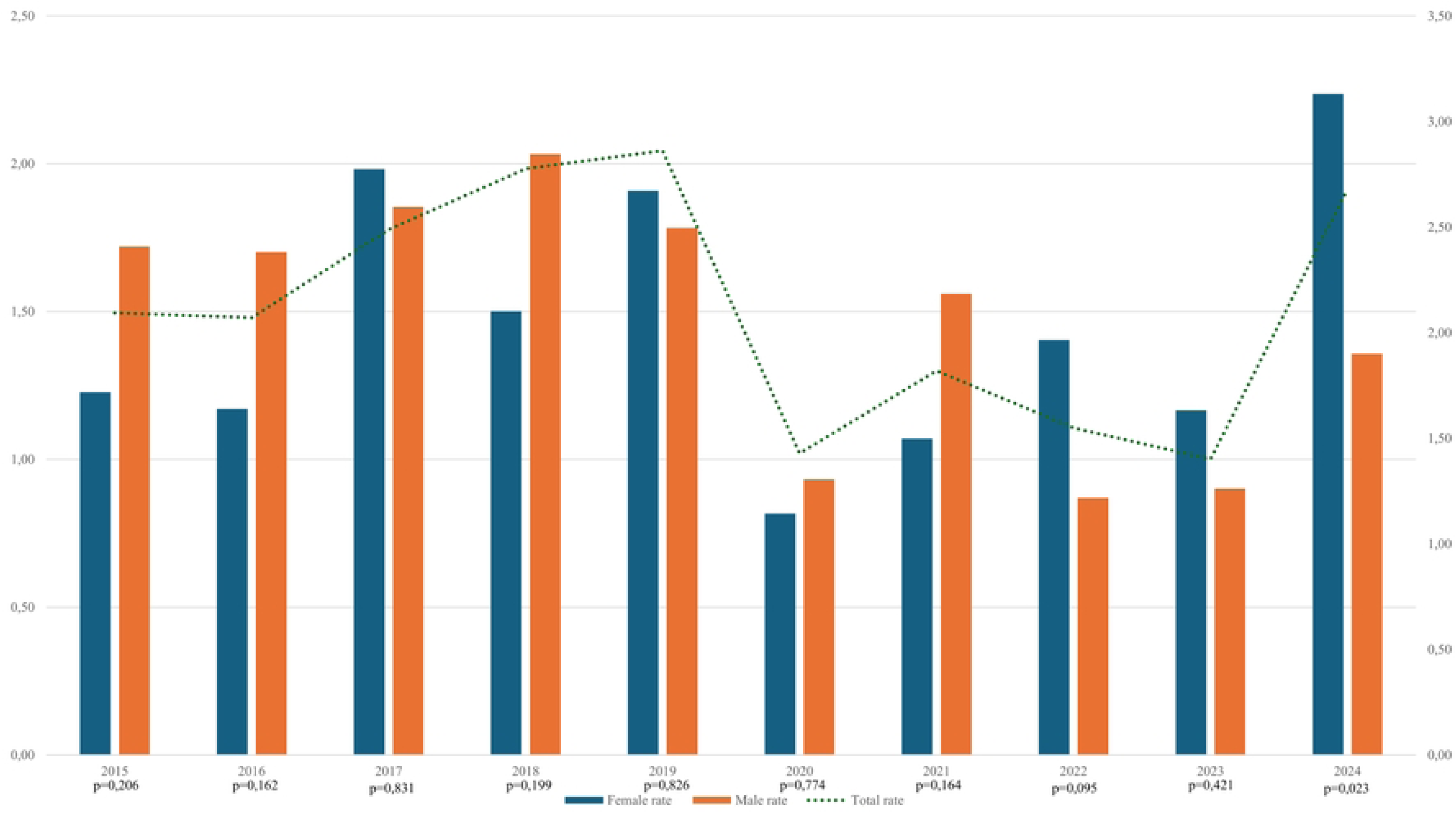
Distribution of the prevalence rate by sex of textiloma in Colombia for the period 2015– 2024.

In 2020 and 2021, male rates were higher, while in 2022, 2023 and 2024 female prevalence was higher. When comparing male and female rates, statistically significant differences were found in 2024 (p=0.023), a year in which there were more female cases (N=89) than male cases (N=52). In relation to the total rate, there was a progressive increase until 2019, followed by a reduction in 2020 and a new increase in 2024 with a rate = 2.68 (N=141).

The health insurance system analysis shows that the distribution of cases across time depends on the type of regime (p=0.000). During the study period, the highest number of cases corresponded to the contributory and subsidized regimes. In the contributory regime, notable increases were observed in 2018 (N=64), 2019 (N=70), and 2024 (N=86), the year with the highest number in the period. In the subsidized regime, cases remained relatively stable between 2015 and 2019 (N=57-61) but descended to 38 in 2020 with subsequent variations in 2023 and 2024 (N=27 and N=54, respectively). When comparing both systems, in most years there were no statistically significant differences, except in 2024, where the difference between the contributory (N=86) and subsidized (N=54) systems was notable. (Table 2)

**Table 1.**
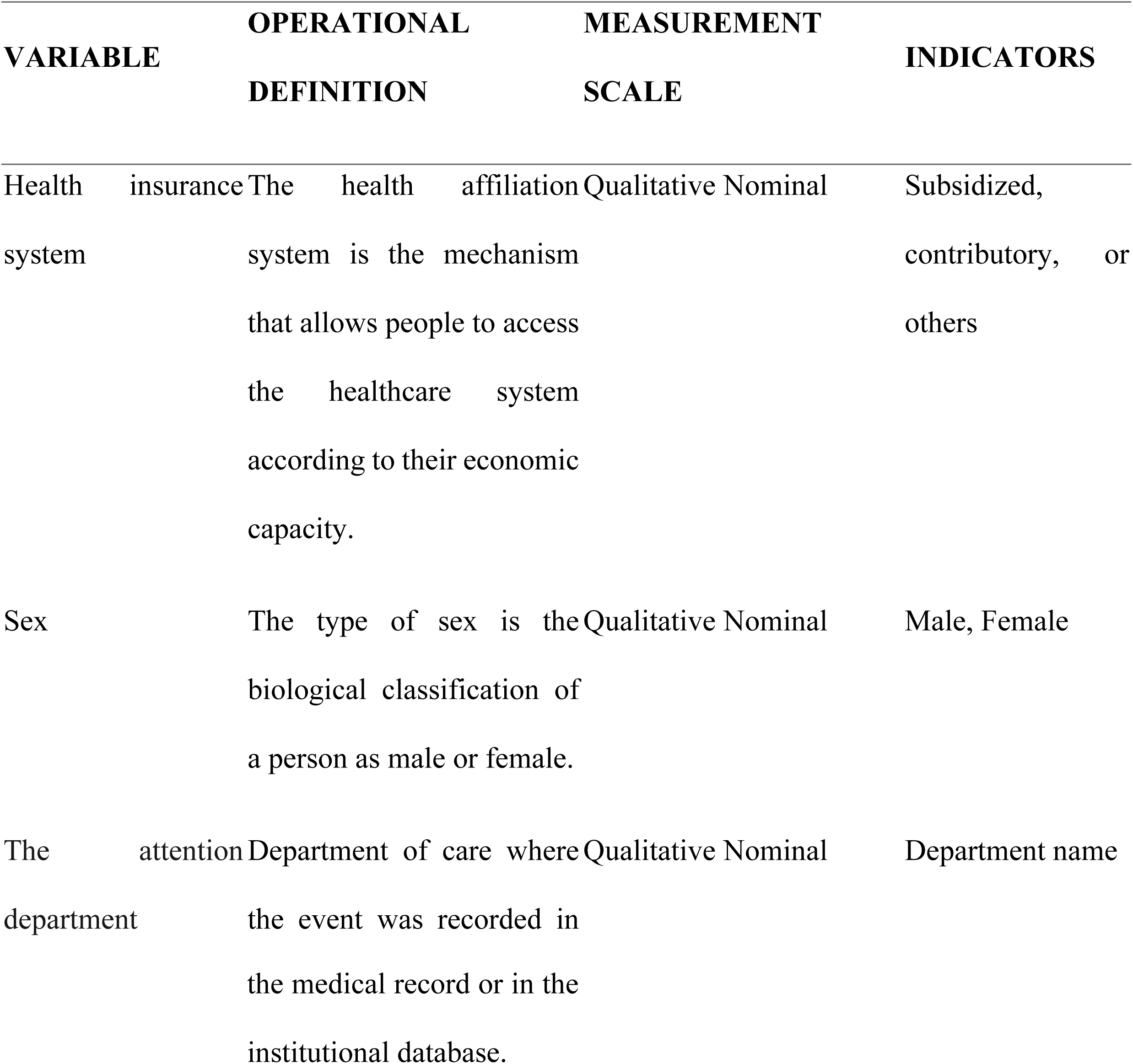
Operationalization of study variables.

**Table 2.**
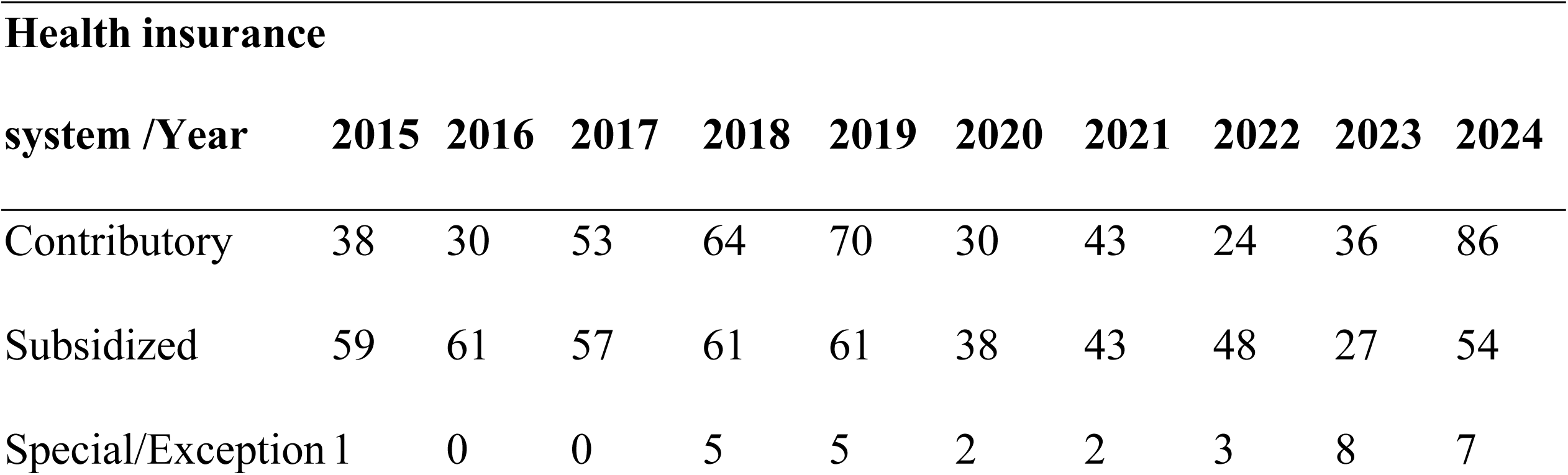
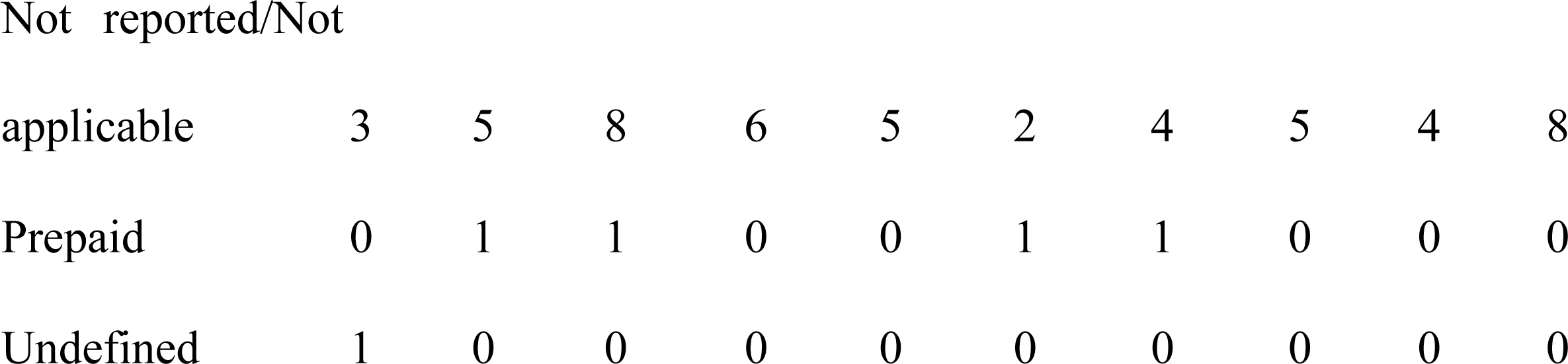
Distribution of textiloma workers by health insurance system in Colombia during the period 2015–2024.

When analyzing the average rate for the period, it was found that the departments with the highest rates were Putumayo with 5.76±4.40 [2.59-14.67], followed by Magdalena with an average of 4.13±2.16 [1.97-8.40], then Quindio with 3.85±2.21 [1.80-7.41], followed by Meta with 3.55±2.29 [0.98-9.41], and finally Boyaca with 3.20±2.62 [0.80-8.57]. The departments with the lowest rates were Guainia, Vaupes, and Vichada with 0.00±0.00 [0.00-0.00], followed by La Guajira with 0.31±0.47 [0.00-1.09] and Nort of Santander with 1.10±1.15 [0.60-4.02]. (Figure 3).

**Figure 3.**
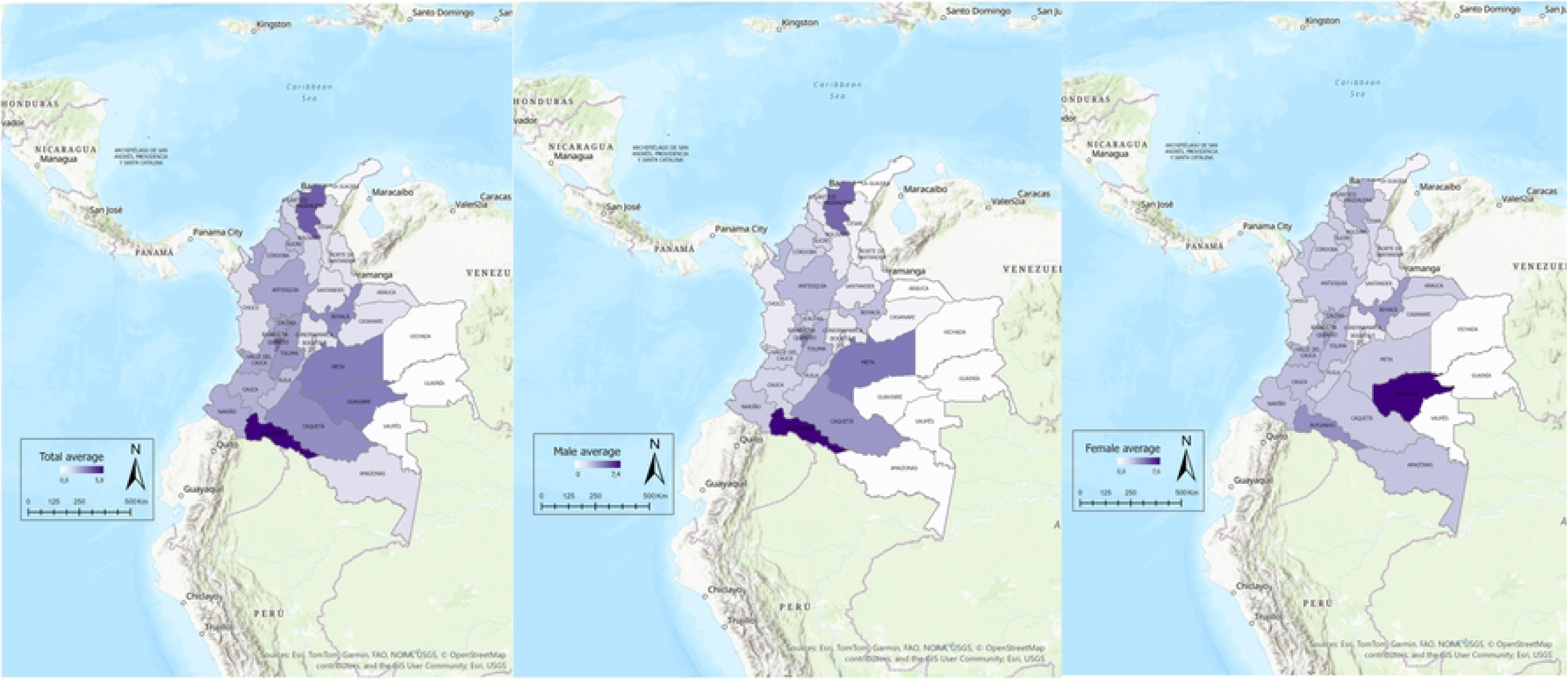
Geographic distribution of the incidence rate of textilomas 2015-2024, by department in cases per million inhabitants.

It was found that, by sex, men had higher averages than women in the departments of Putumayo, where men registered 7.41±5.86 [2.59-17.72], while women reached 4.10±7.04 [0.00-23.76]. In Magdalena, men had 5.12±4.19 [1.97-13.71], compared to 3.14±1.83 [0.00-7.45] in women. Similarly, in Meta, men reported 4.67±4.29 [0.98-14.89], while women reported 2.40±1.66 [0.00-5.57]. In Quindio, averages of 4.15±5.17 [0.00-14.94] were observed for men and 3.56±1.58 [0.00-7.05] for women. However, there were notable exceptions in the Archipelago of San Andres, Providencia and Santa Catalina, where the average for women was 6.17 ± 18.51 [0.00-31.62] and in Guaviare 7.56 ± 16.13 [0.00-25.28]. However, there was no report for the male population.

The male standard deviation shows relatively homogeneous values, with cyphers below 2 in most departments, although there are also some significant values in Putumayo with SD=5.86, CV=79% and Quindio with SD=5.17, CV=125%, indicating heterogeneity in these regions compared to the others in the time trend.

In the female population, the variability of SD is more relevant, with cases such as the Archipelago of San Andres, Providencia, and Santa Catalina SD=18.51, CV=300% and Guaviare with SD=16.13, CV=213%, which contrast sharply with other departments where dispersion is minimal, such as Antioquia with SD=1.47, CV=59% or Cundinamarca with SD=0.86, CV=74%. This shows that in the female group, the distribution of data is the most heterogeneous.

Finally, in the overall total, the standard deviation of the departments remains between 1 and 3, although again there are outliers such as San Andres with SD=9.49, CV=300%, Guaviare with SD=7.58, CV=213% and Putumayo with SD=4.40, CV=76%.

In relation to the analysis of department by year rates, overall notable heterogeneity was observed across regions, with some departments showing wide interannual fluctuations and others exhibiting consistently low values.

Putumayo showed the greatest variability over the series, with the total range extending from 0.00 to 14.67 million inhabitants. However, the largest outlier in the distributions corresponds to the Archipelago of San Andres, Providencia and Santa Catalina in 2024 (value ≈ 31.62), suggesting a singular peak that requires detailed verification. Magdalena and Quindio also displayed moderately high dispersion (Magdalena [1.97 - 8.40]; Quindio [1.80 - 7.41]). That indicates episodes with notable increases in certain years.

Regarding measures of central tendency, Putumayo recorded the highest median (med = 4.19) million inhabitants, followed by Magdalena (med = 3.65) million inhabitants and Tolima (med = 3.02) million inhabitants. By contrast, Amazonas, Guainia, Guaviare, Vaupes and Vichada showed the lowest medians (med ≈ 0) million inhabitants and very small boxes, it may reflect stability at low prevalence levels throughout the decade. (Figure 4)

**Figure 4.**
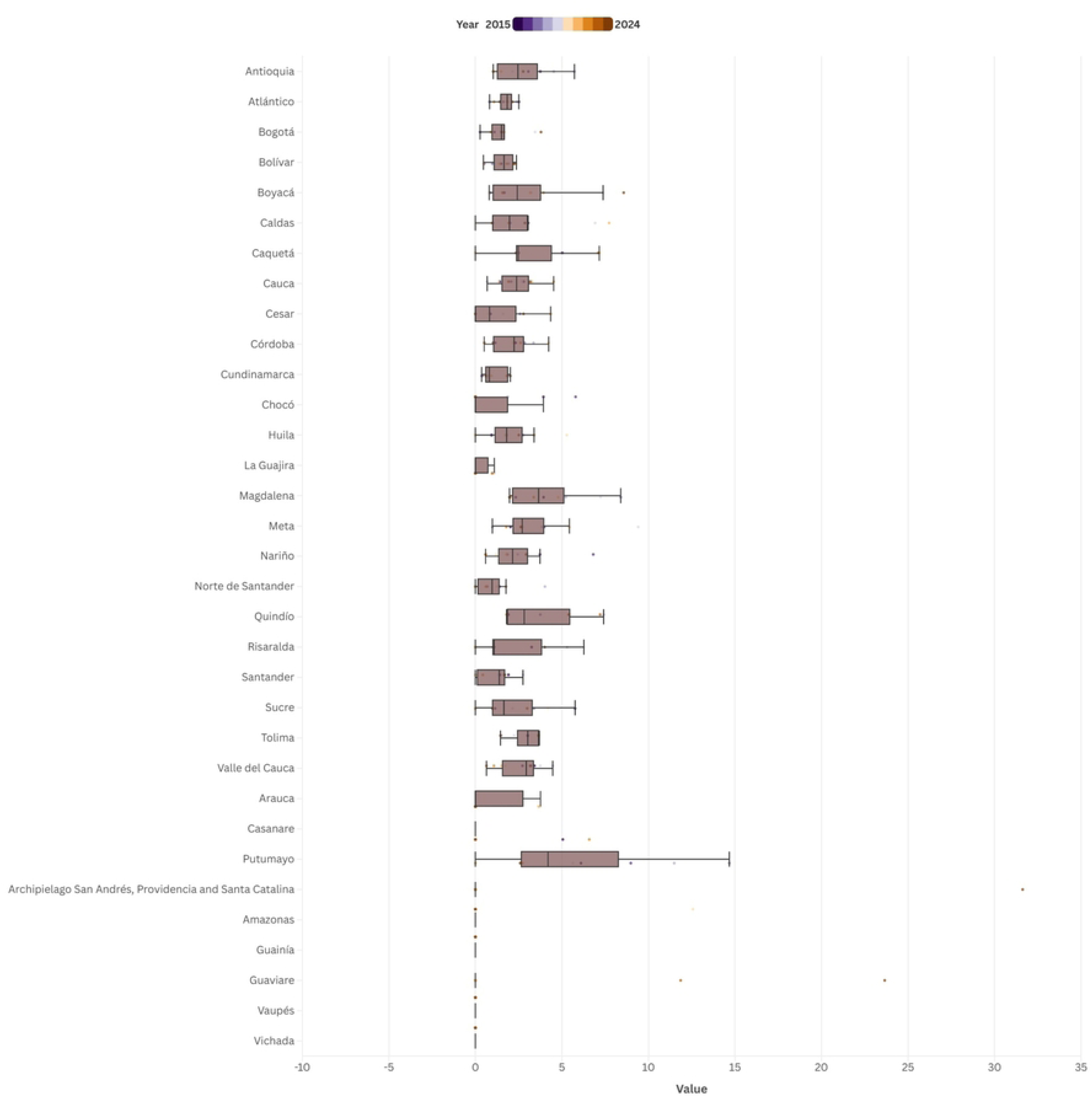
Distribution rates of textilomas by department and year, Colombia, 2015–2024.

When observing the distribution of men rates by department on an annual basis, regional differences in the prevalence of textilomas were identified during the study period.

Quindio showed the greatest variability and the greatest interannual dispersion in the series, with a total range of 0.00 to 14.94 (med = 1.89) million inhabitants. The width of the box and whiskers suggests significant fluctuations between years and the possibility of specific episodes with increases in the rate. Magdalena also showed high variability, with a total range of 0.00 to 13.71 (med = 3.81) million inhabitants; both departments recorded episodes with notable increases in specific years, although no extreme outliers were detected.

Putumayo showed high variability and the most notable extreme outlier in the series: an outlier in 2016 with a rate of approximately 17.72 million inhabitants. The total range observed in Putumayo was 0.00 to 17.72 (median = 5.66), highlighting specific episodes of notable magnitude.

In contrast, the departments of La Guajira, Archipelago of San Andres, Providencia and Santa Catalina, Arauca, Casanare, Amazonas, Guainia, Guaviare, Vaupes, and Vichada showed medians close to zero and low dispersion throughout the decade, reflecting stability at low prevalence levels. (Figure 5).

**Figure 5.**
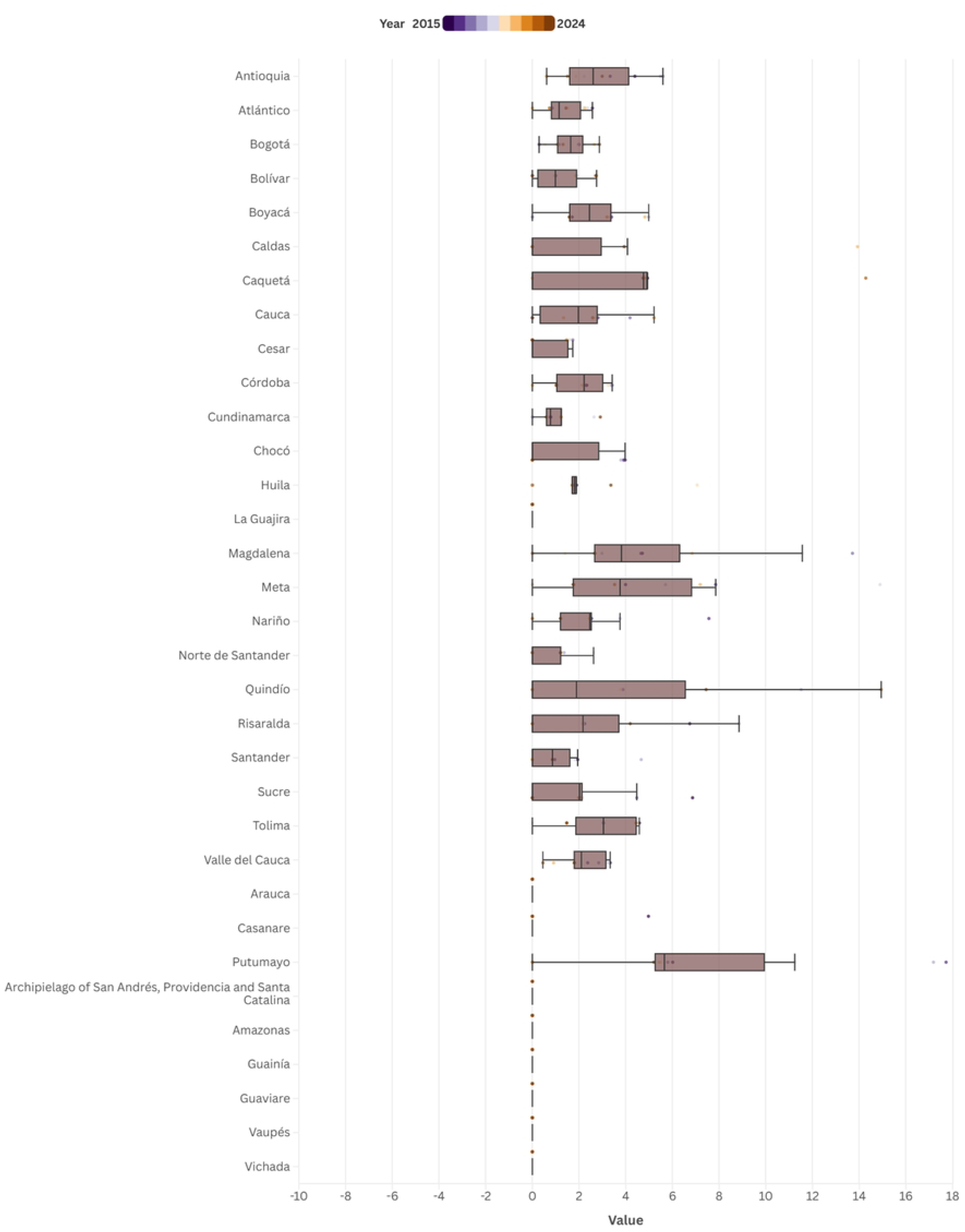
Distribution men rates of textilomas by department and year, Colombia, 2015–2024.

When examining the distribution of women rates by department on an annual basis, clear regional differences in the prevalence of textilomas were identified during the study period.

Boyaca showed the greatest variability, with the highest interannual dispersion in the series, ranging from 0.00 to 15.41 (med = 1.57) million inhabitants. The width of the box and whiskers suggest significant fluctuations throughout the period. Similarly, Caqueta showed high variability, with a range of 0.00 to 9.53 (med = 0.00) million inhabitants.

The largest outlier corresponds to the Archipelago of San Andres, Providencia and Santa Catalina in 2024, with a rate of ≈ 61.70 million inhabitants, followed by Guaviare in 2024, with a rate of ≈ 50.34 million inhabitants, values markedly higher than the rest of the observations. The department with the highest median was Quindio (med = 3.55) million inhabitants, followed by Tolima (med = 2.91) million inhabitants, indicating that, in terms of central tendency, these territories maintain rates higher than most of the country.

The regions with the lowest dispersion include Casanare, Amazonas, Guainia, Vaupes and Vichada, which had medians close to zero and low variability throughout the decade, reflecting stability at low prevalence levels. (Figure 6)

**Figure 6.**
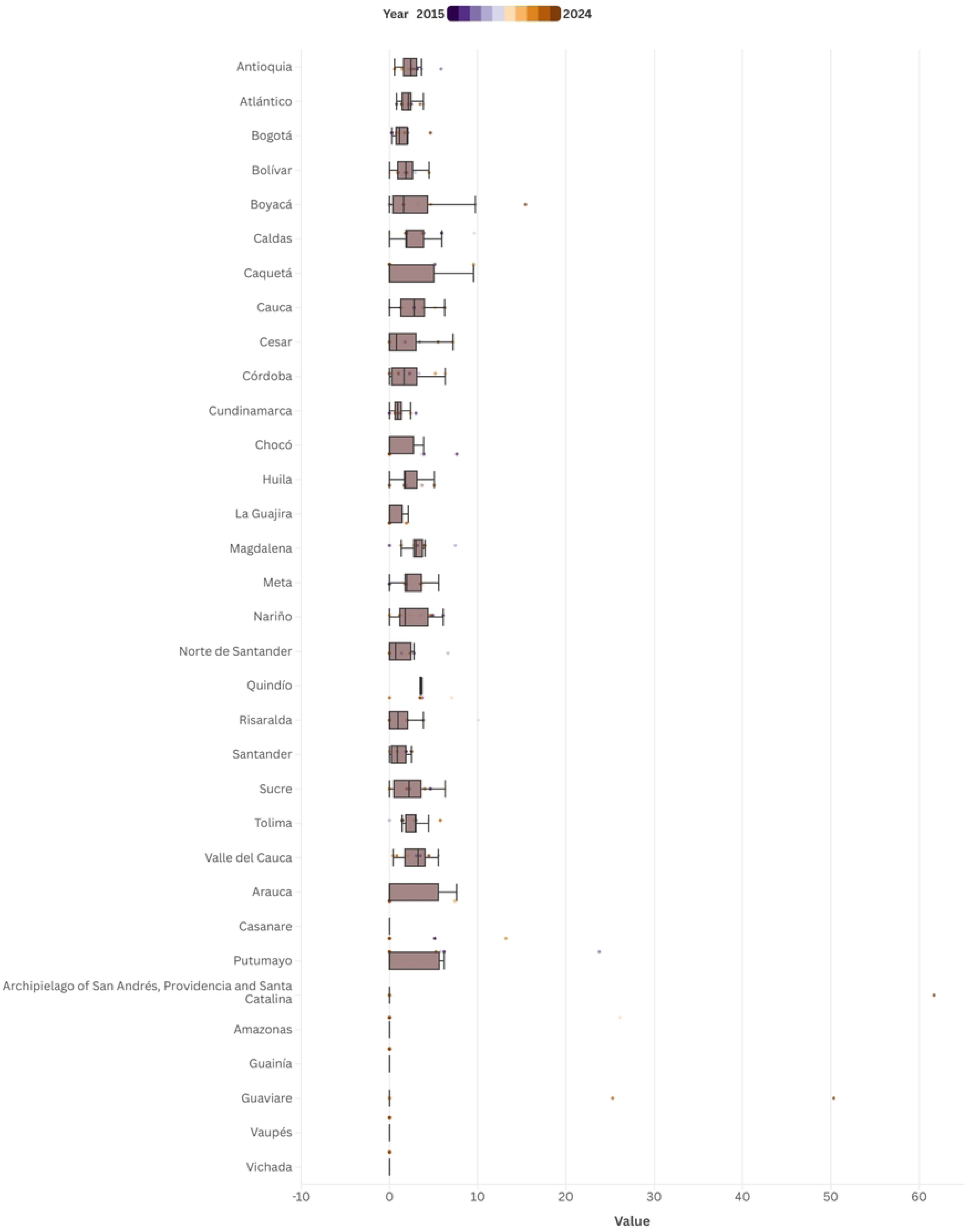
Distribution rates of textilomas by department and year in women, Colombia, 2015–2024.

To represent a comparative point of view related between departments, we have designed a ranking to show the variations of the textiloma rate among the years 2015, 2019, and 2024.

The departments that remain at the top of the rankings show consistent behavior, albeit with high rates, which is concerning. In the case of Valle del Cauca, it ranked 9^th^ in 2015 with a rate of (3.41), rising to 7^th^ place in 2019 with a rate of (3.74), a position maintained through 2024.

Meanwhile, the department of Nariño remained stable throughout the three periods analyzed, ranking 8^th^, 10^th^, and 9^th^, with an average rate between (3.72 - 2.93). Finally, Risaralda moved from 10^th^ place in 2015 to 5^th^ in 2024, with a rate of (4.00).

On the other hand, some departments exhibited more heterogeneous behavior. In the case of Arauca, in 2015 it ranked 25^th^ with an incidence rate of ≈0, rising to sixth place in 2019 with a rate of (3.75), and falling again to 26^th^ place in 2024, with a rate of ≈0. Similarly, the department of Tolima ranked 12^th^ in 2015, with a rate of (3.02), and by 2024 it had fallen significantly to 22^nd^ place, with a lower rate of (1.44). The departments that do not show any performance in the graph correspond to those for which no data was recorded during those years. (Figure 7)

**Figure 7.**
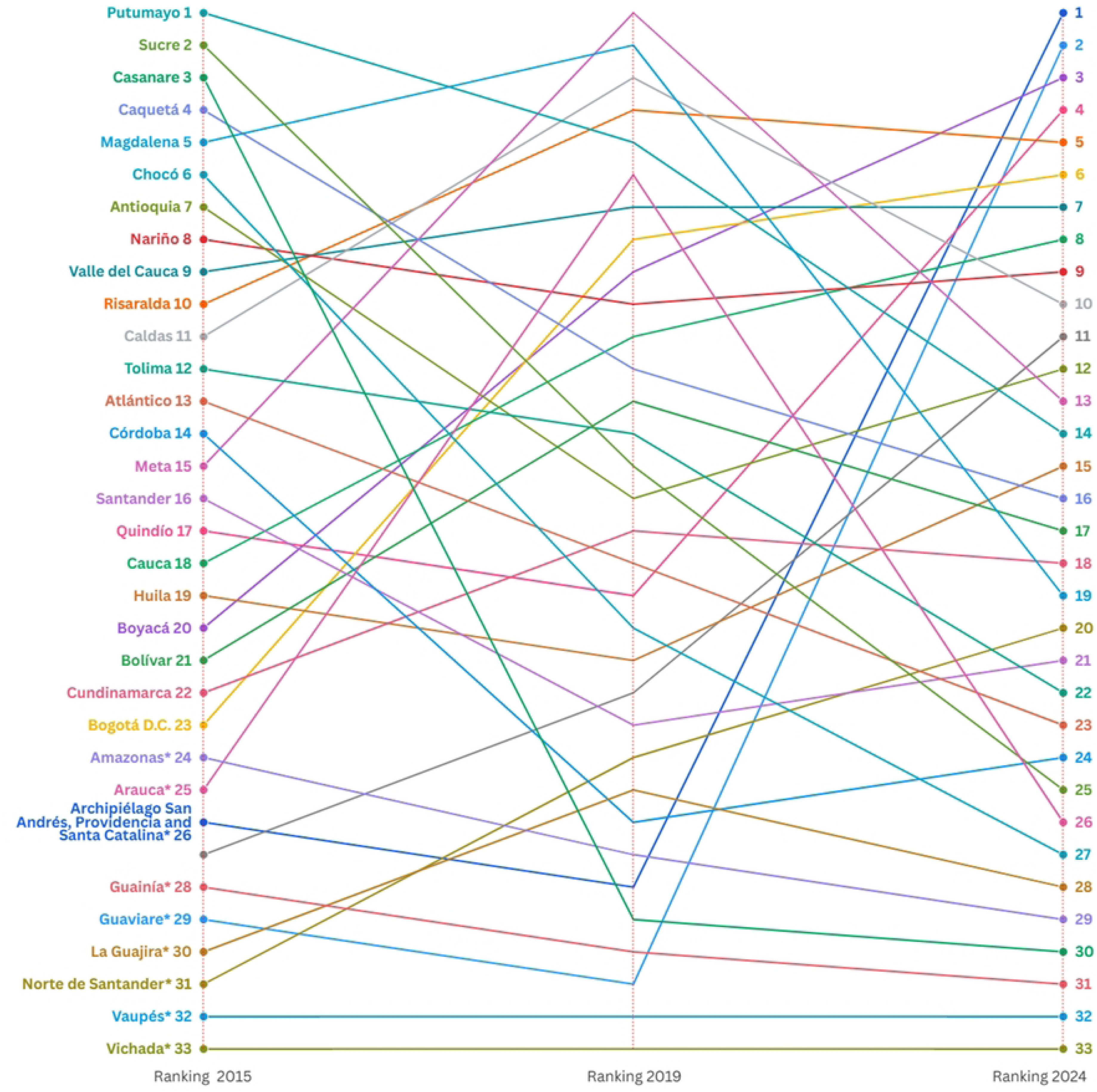
Rates ranking for the years 2014, 2019, and 2024.

## Discussion

According to the data presented, the behavior of the rates between new and repeated cases during the period 2015–2024 is heterogeneous, as more new cases (1,446) were reported compared to repeated cases (646). Between 2015 and 2019, there is evidence of a steady increase in the total number of reported cases, unlike the new cases, which show a more stable pattern between 2016 and 2019.

Related to the textiloma rates by sex during the period 2015-2024, a uniform pattern is identified in most years, with no statistically significant differences between men and women distributions. This result indicates that the distribution of cases has remained balanced between the two groups. However, in 2024, there is a notable variation from the previous pattern, showing a significant difference (p=0.023) associated with a marked increase in the female rate (2.24) compared to the male rate (1.36).

When analyzing the distribution of textiloma cases by health insurance regime between 2015 and 2024, a dependent relationship between the type of regime and the study years becomes evident. In the early period (2015–2016), notable gaps were observed between the contributory and subsidized systems, with a difference of 21 cases and a higher concentration in the subsidized system. Subsequently, the gap between the two regimes tended to narrow, showing some fluctuations in the number of cases. However, by 2022, a difference of 24 cases was observed, again favoring the subsidized regime, and in 2024, the disparity reappeared, with a marked increase in cases within the contributory regime, reaching a difference of 32 cases in the final year. It may provide an interesting overview of inequalities across regimes or disbalanced behavior in the surgical procedures by the timeline.

The analysis of the total average, calculated per million inhabitants, showed that the highest values were again located in Putumayo and San Andres, while the eastern departments recorded the lowest averages. The total standard deviation remained between 1–3, although outlier values persisted in San Andres (SD=9.49; VC=300%), Guaviare (SD=7.58; VC=213%), and Putumayo (SD=4.40; VC=76%). That is, the highest values were concentrated in the southern and northern departments, while the eastern departments showed the lowest averages.

In the male group, the departments with the highest average values were Putumayo (7.41), Magdalena (5.12), Meta (4.67), and Quindio (4.15), which suggests a concentration of high values in specific areas of the south and center of the country. The standard deviation showed a relatively homogeneous distribution, with values ≈2 in most departments, although Putumayo (SD=5.86; VC=79%) and Quindio (SD=5.17; VC=125%) stood out, where the dispersion was considerably higher, showing a more variable temporal trend compared to the other regions.

In contrast, the female population presented a more dispersed spatial distribution and greater internal variability. Although the averages were lower than those of males in most departments, relevant exceptions were identified in Guaviare (7.56) and the Archipelago of San Andres, Providencia and Santa Catalina (6.17), where no male values were reported. These areas also showed the highest heterogeneity, with extremely high standard deviations (SD=16.13; VC=213% and SD=18.51; VC=300%, respectively).

When analyzing the distribution of rates by department between 2015 and 2024, a marked regional variability in the prevalence rate of textilomas was evident. Putumayo exhibited the greatest interannual dispersion, with a range of approximately 0.00 to 14.67 (med = 4.19) million inhabitants, reflecting wide fluctuations in the occurrence of cases over time. Furthermore, the most extreme outlier corresponded to the Archipelago of San Andres, Providencia, and Santa Catalina, which recorded a rate close to 31.62 million inhabitants in 2024, a value that significantly deviates from the pattern observed in the rest of the country. In contrast, several departments showed low medians (≈0), indicating stability at low prevalence levels throughout the study period.

In relation with them distribution of men rates per million inhabitants by department during the 2015–2024 period, marked heterogeneity in data dispersion was evident. Quindio, Magdalena, and Putumayo showed the greatest interannual variability, with wide ranges reflecting considerable fluctuations in the occurrence of cases. In particular, Putumayo recorded the widest range of values (0.00–17.72; med= 5.66) million inhabitants, along with the most extreme outlier, suggesting notable instability in the prevalence of the event in this area. Similarly, Quindío and Magdalena exhibited high dispersion, with medians of approximately 1.89 and 3.81 million inhabitants, respectively. In contrast, about one-third of the departments maintained low rates and limited variability, indicating stability in the trend throughout the study period.

In the analysis of the distribution of woman rates by department during the period 2015– 2024, a marked heterogeneity in data variability was evident. The departments of Boyaca and Caqueta showed the greatest interannual dispersion, with wide ranges reflecting significant fluctuations in the occurrence of cases over time. In particular, Caqueta exhibited considerable dispersion, with values reaching up to 9.53 million inhabitants, while Boyaca displayed wide variability, with values as high as 15.41 million inhabitants and the presence of outliers in 2024.

Meanwhile, the Archipelago of San Andres, Providencia, and Santa Catalina recorded the most extreme outlier, with a value of 61.70 million inhabitants, representing a substantial deviation from the pattern observed in the rest of the departments. In contrast, the departments of Casanare, Amazonas, Guainia, Vaupes, and Vichada maintained medians close to zero and reduced variability, suggesting stability in the rates throughout the period analyzed.

A more detailed analysis reveals notable variations in the rankings among the different departments, demonstrating heterogeneous behavior. For example, Meta ranked 15^th^ in 2015, rose to 6^th^ in 2019, and then fell back to 13^th^ in the most recent period, with a rate of (2.62). The department of Sucre shows a downward trend: in 2015 it ranked second with a rate of (5.76), while in 2019 it dropped to 25^th^ position, with a rate of (0.98).

In the case of San Andres, a concerning trend is evident, as it ranked 26^th^ in 2015 with a rate of (0), and in 2019 it abruptly rose to 1^st^ place, with a rate of (31.62) Similarly, Guaviare experienced significant fluctuations in 2015 it ranked 29^th^, descended to 31^th^ that same year, and subsequently rose to second place, with a rate of (23.65). Finally, the department of Choc0 ranked 6^th^ in 2015 with a rate of (3.92), fell to 20^th^ in 2019 with a rate of (1.83), and by 2024 it had fallen to 27^th^ place, with (0) rate.

The underreporting of textiloma cases represents a critical barrier to effective surveillance and the improvement of surgical safety in Colombia. Many of these adverse events are not systematically documented due to multiple limitations: the absence of specific protocols for reporting them, a lack of education and awareness about their notification, and fear of sanctions or legal consequences. This situation prevents an accurate understanding of the behavior of textiloma, hinders the analysis of their causes, the implementation of preventive strategies of measurement, and the collection of reliable data necessary to design interventions that reduce this issue. Consequently, the official reports may underestimate the true magnitude of the phenomenon, compromising both transparency and patient safety (10).

An important limitation of this study lies in the fact that cases were attributed to the location where the textiloma was diagnosed, rather than to the site where the surgical intervention that originated was performed. This approach may introduce a location bias, as the place of detection does not necessarily correspond to the site of the initial surgical event. Given the natural history of this condition, often characterized by delayed and nonspecific presentation, it is likely that a considerable interval elapses between the surgical procedure, the onset of symptoms, and the definitive diagnosis, then the treatment that is the origin of these reports. During this period, patients may have changed their place of residence or received care in healthcare institutions different from those where the original surgery was conducted. Consequently, geographic assignment based on diagnosis may not accurately reflect the true origin of the event, thereby limiting the territorial interpretation of the results and the identification of potential institutional or regional patterns associated with the risk of occurrence.

In the Colombian context the public policy related with patient safety establish a regulatory framework applicable to the prevention of textilomas through the promotion of surgical safety, infection control, and the surveillance of adverse events. That corresponds to the international framework promoted by multilateral institutions like the WHO and PAHO. WHO (The adoption of the World Health Organization) Surgical Safety Checklist in Colombian hospitals has been one of the most significant advances in reducing errors. This tool, when applied rigorously, allows for confirmation of the patient’s identity, the correct surgical site, and the complete count of instruments and materials before closing the surgical wound (12).

Among the existing regulatory documents, Resolution 1441 of 2013, issued by the Ministry of Health and Social Protection, defines the conditions for the authorization of healthcare service providers, including quality standards, patient safety requirements, and the operation of surgical services. It mandates the implementation of asepsis protocols, surgical safety checklists, and instrument control procedures, all essential measures for the prevention of textilomas (13).

Resolution 3100 of 2019 updated healthcare service authorization standards, reinforcing biosafety, sterilization, and material management protocols to reduce the likelihood of surgical incidents, including textilomas (14). Then, the Patient Safety Policy promotes a culture of safety in healthcare institutions, encouraging identification, reporting, and analysis of adverse events. Likewise, External Circular 030 of 2014 of the National Health Superintendence, a control, inspection and vigilance institution, establishes guidelines for risk management in healthcare and the reporting of adverse events (15).

Then, during the COVID-19 pandemic, a marked decrease in surgical activity was observed nationwide (16). Several reports indicated that between 2020 and 2021 there was a substantial reduction in the number of procedures, both elective and emergency, accompanied by a reorganization in hospital service delivery and in the operational capacity of operating rooms. This trend occurred in the context of medical care prioritization and temporary adjustments in health service provision, which temporarily altered the usual dynamics of the surgical system. Therefore, the period corresponding to the pandemic can be regarded as a turning point within the analyzed timeframe, as it reflects a transient variation in surgical care patterns in Colombia, widely documented in national literature (17), this context can explain the data distribution in some lower points in the trend.

The regional distribution of textiloma cases in Colombia reveals a significant territorial gap, with a higher concentration in departments such as Valle del Cauca, Nariño, Risaralda, Meta, and Chocó. The monitoring and documentation of surgical events are essential for controlling this occurrence. In many cases, patients from rural or low-resource municipalities opt to turn to neighboring departments to receive specialized medical care, due to the lack of well-equipped hospitals or trained personnel in their places of origin. This creates a heavier workload for healthcare centers in these areas and may contribute to an increase in newly reported cases in other departments. Furthermore, differences in infrastructure, human resources, and the availability of information systems across regions affect the ability to detect, report, and adequately monitor adverse surgical events. Thus, the regional gap reflects not only a geographical pattern but also inequalities in access and in the health system’s capacity to respond to textiloma cases in the country (18).

Preventing textilomas requires more than merely complying with a protocol; it also involves strengthening human and organizational capacities within the health system. A strategy to strengthen the prevention of textilomas in Colombia could be the implementation of a national intelligent system for surgical analysis and traceability, capable of integrating data collected during each procedure (counts, surgical times, and outcomes). This system would enable the generation of automatic indicators and risk maps by department or institution, facilitating evidence-based decision-making for the design of effective public policies.

The creation of a culture of shared responsibility, in which each team member understands that preventing a textiloma depends on attention to detail, assertive communication, and adherence to protocols, is essential. The existing regulatory mechanisms in Colombia provide a foundation for the prevention of these events, but their effectiveness depends on their implementation within institutions.

About it, in the regional context, countries like Venezuela have been implemented strategies to prevent retained surgical items in surgical procedures (9). These include conducting a thorough count and custody of all materials entering the surgical field in each operative act, as well as the obligation for the surgical technologist to document in the medical record the quantity and quality of materials used or unused during the procedure (9). Additionally, it is required to promptly report any inadequate condition of surgical materials to the corresponding health institutions and to carefully review the notes of other healthcare professionals, especially nursing assistants, ensuring they are consistent with the records and the of surgical instrumentist or technicians to improve an multidisciplinary view across the patient safety policies.

### Conclusion

In conclusion, the behavior of textiloma cases in Colombia between 2015 and 2024 reveals a heterogeneous distribution influenced by territorial, institutional, and structural factors within the health system. The persistence of this adverse event, despite existing regulations and protocols, underscores the need to strengthen surgical safety practices and reporting mechanisms. Likewise, enhancing information systems, ensuring the traceability of surgical materials, and implementing technological strategies that guarantee the safe counting of instruments are priority actions to reduce the incidence of these preventable events. Finally, consolidating a patient safety policy grounded in shared responsibility, transparency, and continuous improvement will foster progress toward a safer, more equitable, and quality-oriented healthcare system in Colombia.

## Data Availability

The data is available at Mendeley Data 10.17632/fr594gjr4c.1

